# Real-time hip biomechanics from smart garments via a physics-informed neural network

**DOI:** 10.64898/2026.05.06.26352104

**Authors:** Bradley M. Cornish, Claudio Pizzolato, David J. Saxby, Nathan R. Lyons, Yana A. Salchak, Matthew T.O. Worsey, David G. Lloyd, Laura E. Diamond

## Abstract

Tissue-level mechanical stimuli are primary drivers of tissue adaptation and can be optimised during conservative treatments to improve treatment outcomes for many highly prevalent musculoskeletal conditions. Current laboratory-based technologies limit our ability to connect conservative interventions such as exercise and movement modification with muscle, joint, and tissue-level mechanics, in natural environments. We introduce a physics-informed neural network (PINN) to estimate clinically relevant biomechanics from smart garments. By accounting for physiological dynamics of neural activation and muscle contraction, the PINN accurately predicted hip joint angles (RMSE <6 degrees), moments (RMSE 0.12 N·m/kg to 0.30 N·m/kg), and joint forces (RMSE 6 to 16%) from three inertial measurement units and four electromyographic sensors. We demonstrated that the trained PINN can be combined with a smart garment to estimate hip biomechanics, in real-time, during a gait retraining intervention aimed at modifying joint loading to treat hip osteoarthritis. The developed PINN and smart garment system may be adapted and generalised for personalised management or rehabilitation of a broad range of musculoskeletal diseases and injuries, in clinical, home, workplace, and sporting environments.

## Introduction

Musculoskeletal conditions are major causes of personal and societal burden^1^. Globally, rates of musculoskeletal conditions are increasing^2^, and healthcare costs are the highest on record and continue to rise^3^. Conservative interventions (e.g., physical activity and exercise) are globally recommended as first-line treatments for musculoskeletal conditions^4^. However, conservative interventions have limited efficacy for many highly prevalent conditions such as osteoarthritis^5^ and tendinopathy^6^. These limited treatment effects can be explained by our poor understanding of how conservative interventions influence musculoskeletal tissue state through mechanical stimuli^7^. When mechanical loading is applied to musculoskeletal tissues, the resulting stresses and strains drive musculoskeletal tissue adaptation and are important biomarkers of tissue health^8-11^. Yet, we remain unable to directly connect individualised physical activity and exercise (e.g., type, intensity, duration) with tissue and cellular level mechanobiological responses to modify disease state^12,13^. For instance, dynamically cyclical articular loading compresses and decompresses cartilage, creating cyclic fluid flow within and strains the collagen matrix, the interlinked chondrocytes, and other molecules^14^. This stimulation of the mechanobiological pathways which support cartilage function and drive remodelling offers a potential modifiable treatment target for managing osteoarthritis, i.e., articular load optimisation^9,11^. At present, we lack technology to estimate articular cartilage mechanics and many other mechanical biomarkers of tissue health (e.g., tendon and bone strain) in natural environments, impeding the development of effective therapeutic interventions to treat many highly prevalent and burdensome musculoskeletal conditions^15^.

International research strategies and governing bodies have acknowledged the lack of clinically relevant and cost-effective digital health technologies, and called for increased investment and development^16,17^. Inertial measurement units (IMU) are a promising technology for real-world estimation of mechanical biomarkers^18^, as they enable portable and continuous monitoring of movement in natural environments^19^. However, advances in IMU technology have primarily related to improving accuracy and reliability of their measurements of body motion^20^. Although important to understanding function, body motion alone does not drive tissue adaption^7^, and does not account for the influence of forces external and internal to the body, nor does it account for variations in anatomy and neural drive which influence tissue state^21^. Extending upon the capacity of IMU, wearable smart garments can facilitate concurrent recording of motion and neural activity through a combination of IMU and portable electromyography (EMG)^22^. However, without measuring forces applied externally to the body (e.g., between foot and ground), smart garments have limited compatibility with gold-standard physics-based models to calculate muscle and joint forces. This limitation of smart garments could be circumvented by using artificial neural networks (NN) for rapid estimation of clinically relevant mechanical biomarkers (e.g., joint loads and tissue mechanics)^23^ in ecologically valid environments and at very low cost compared to laboratory-based motion capture analysis^24^. For example, gait retraining has long been proposed as a method for optimising articular cartilage mechanics to manage osteoarthritis^25^ but the inability to estimate joint loading, or forces, in natural environments has hindered efforts to assess treatment efficacy^15^. Real-time estimation of joint forces using a smart garment could see biofeedback-informed gait retraining become a cost-effective therapeutic option for musculoskeletal conditions, like osteoarthritis.

To date, NN have been used to accurately predict joint angles and moments using IMU^26^, to calculate full-body and joint-level kinematics^27^ and Achilles tendon force^28^ using anatomical key point data derived from bi-planar video data, and to estimate hip contact forces from sparse motion data combined with EMG^29^. Despite these notable applications of NN to biomechanical problems, there remains no system for accurate and reliable estimation of joint and tissue loads that is robust under real-world conditions. Poor generalisability and reliability of NN for biomechanical analyses^30^ may be overcome *via* physics-informed neural networks (PINN). A PINN is a NN which integrates physics by penalizing model predictions that violate known physical and/or biophysical laws^31^. For real-world biomechanical analysis, a PINN may be trained to learn the underlying physics and physiology of human motion thus enabling robust estimation of clinically relevant biomarkers of tissue health.

We developed the first smart garment embedded with wearable sensors and a PINN, and trained it using sparse experimental data (i.e., IMU sensors placed on few body segments and EMG recordings of few muscles) to predict biomechanical variables of interest (Fig 1). The PINN embedded the physiological dynamics of neural activation and muscle contraction, and the mechanical properties of force-moment coupling into its network. We then applied the PINN to estimate hip biomechanics (angles, moments, muscle forces, and contact forces) during typical walking and variations (e.g., incline and decline). We assessed validity of the PINN across different human subjects and across a range of motor tasks, a necessity for clinical utility. Additionally, we showed the PINN was responsive to changes in hip contact forces caused by alterations to typical walking. This responsiveness to changes in gait is necessary for the PINN to be used to inform personalised movement modification in the future. Finally, the PINN was combined with a smart garment (with embedded IMU and EMG sensors) for real-time estimation of hip biomechanics outside the laboratory, demonstrating a significant step toward real-world estimation of mechanical biomarkers of tissue health.

**Fig. 1.**
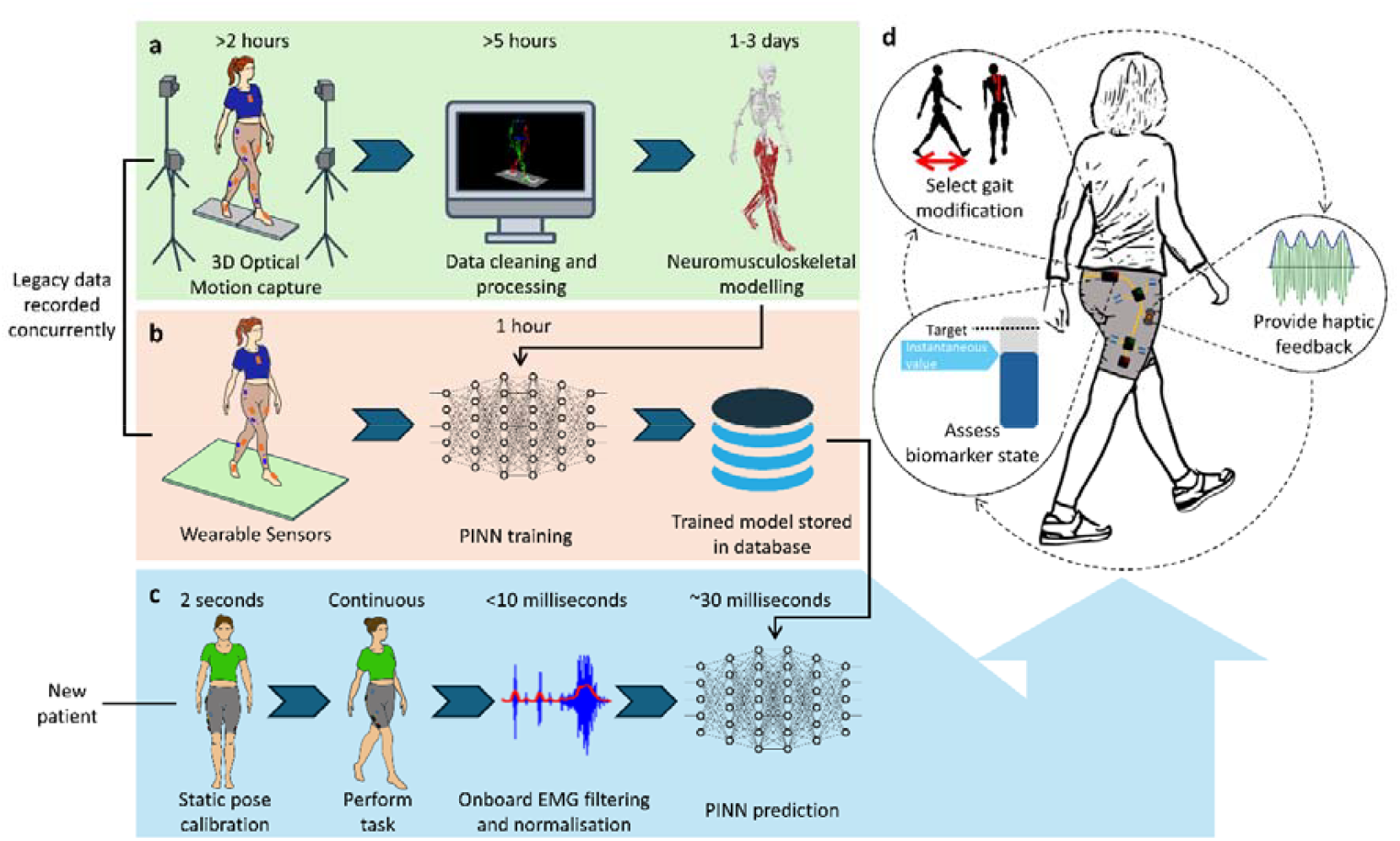
Real-time hip biomechanics computed *via* a physics-informed neural network driven using a smart garment system. **a** Three-dimensional optical motion capture, ground reaction forces, and electromyography (EMG) are recorded in a biomechanics laboratory. Motion capture data are then cleaned, labelled, and filtered *via* a semi-automated process. Laboratory data are then used to drive a neuromusculoskeletal model (NMS) (implemented in OpenSim^32^ and CEINMS^33^) to estimate hip biomechanics. **b** Wearable inertial measurement units (IMU) and EMG are used to train a physics-informed neural network (PINN) where the NMS outputs are used as labelled data. **c** A customised smart garment with three IMU and four EMG sensors is donned by a patient, and a static calibration trial is recorded to determine IMU orientations. The patient then performs a motor task (e.g., walking) and the IMU and EMG data are recorded, processed, and streamed to a trained PINN to estimate hip biomechanics. **d** Real-time operation of the PINN is combined with the customised smart garment to enable biofeedback-assisted gait modification: The PINN estimates a clinically relevant biomarker (e.g., hip contact force), a target is calculated based on subject-specific needs (e.g., increase joint loading), and haptic feedback is provided to prompt the individual to modify their gait.

## Results

### PINN description and architecture

The PINN predicted joint angles (flexion, adduction, rotation), moments (flexion, adduction), 25 muscle forces, and three-dimensional joint contact forces of the hip. To generate training and validation data for the PINN, participants were equipped with marker-based motion capture, full-leg EMG, and lower-body IMU whilst performing multiple movement tasks overground and atop a treadmill with embedded force plates in a biomechanics laboratory. The recorded marker trajectories, ground reaction forces, and EMG were used to calculate lower-body joint angles, joint moments, joint intersegmental forces, muscles forces, and hip contact forces *via* a validated EMG-informed neuromusculoskeletal modelling pipeline (NMS)^32,33^. The PINN was trained to estimate NMS calculated hip biomechanics using 3 IMU and 4 EMG sensors, with participant anthropometrics (i.e., mass, height, body segment dimensions) as inputs. Linear accelerations and angular velocities measured by the smart garment IMU were converted to an anatomical coordinate frame based on a trial of quiet upright stance in anatomical position. These IMU kinematics were combined with EMG linear envelopes normalised to peak values during walking. The IMU and EMG data (100 Hz) were input using overlapping 30-timepoint (300ms) windows.

The PINN was designed to generate physiologically plausible muscle and joint forces at the hip. Previously formulated neural activation^34^ and muscle contraction^35^ dynamics were embedded within, which describe the dynamic relationships between neural drive, muscle-tendon kinematics, and muscle force. To reduce the number of EMG required, synergistic relationships between muscle excitations were leveraged to accurately synthesise non-recorded excitations. As traditionally derived muscle synergies are task specific^36^, we used an autoencoder to generate 25 muscle excitations from 4 measured EMG, with IMU linear accelerations and angular velocities added to the autoencoder input to provide task context. The generated muscle excitations were also penalised to reduce total muscle activation, ensure physiological frequencies (up to 6 Hz), and encourage temporal alignment with measured EMG for muscles with direct mapping (e.g., gluteus muscles). To calculate muscle force, the excitations were input to a Hill-type muscle model^35^ with force-length and force-velocity relationships. We integrated a previously validated NN method to estimate muscle-tendon kinematics^37^, which provided instantaneous muscle-tendon lengths, velocities, moment arms, and lines of action of the relevant muscle about the pelvis. Joint moments were calculated as the sum of the muscle forces with their respective moment arms at each timepoint. Finally, hip contact force was calculated as the sum of NN estimated intersegmental forces with the vector sum of muscles forces collinear to their lines of action on the pelvis.

### Leave-one-subject-out cross-validation

To ensure our PINN generalises to new users, for whom there are no training data, we performed leave-one-subject-out cross-validation. For cross-validation, the PINN predicted highly accurate sagittal plane hip angles, moments, and hip contact force across all tasks (Fig 2). Median root-mean-square-errors (RMSE) for hip angles ranged 3° to 6° across tasks. The hip angles were physiological (i.e., smooth) and without gyroscopic drift common to IMU sensor-fusion based methods for joint angle calculation during continuous movements^24^. Median joint moment RMSE ranged from 0.12 N.m/kg during walking to 0.30 N.m/kg for running. For hip contact force predictions, the RMSE was normalised to the peak hip contact force for each trial. The PINN predicted hip contact force with median RMSE from 6 to 16%. The PINN had the lowest errors for walking (<7%), despite large variations in ground-truth values (range 3 to 6 bodyweight) across different walking speeds and inclines. The low errors of the PINN predictions from sparse wearable data suggest the framework is a viable alternative to expensive laboratory-based systems that rely on slow and technical NMS analysis. Importantly our findings suggest a high level of generalisability across people and tasks, a characteristic that is key to commercial viability, and adoption in the clinic, home, or sport field.

**Fig. 2.**
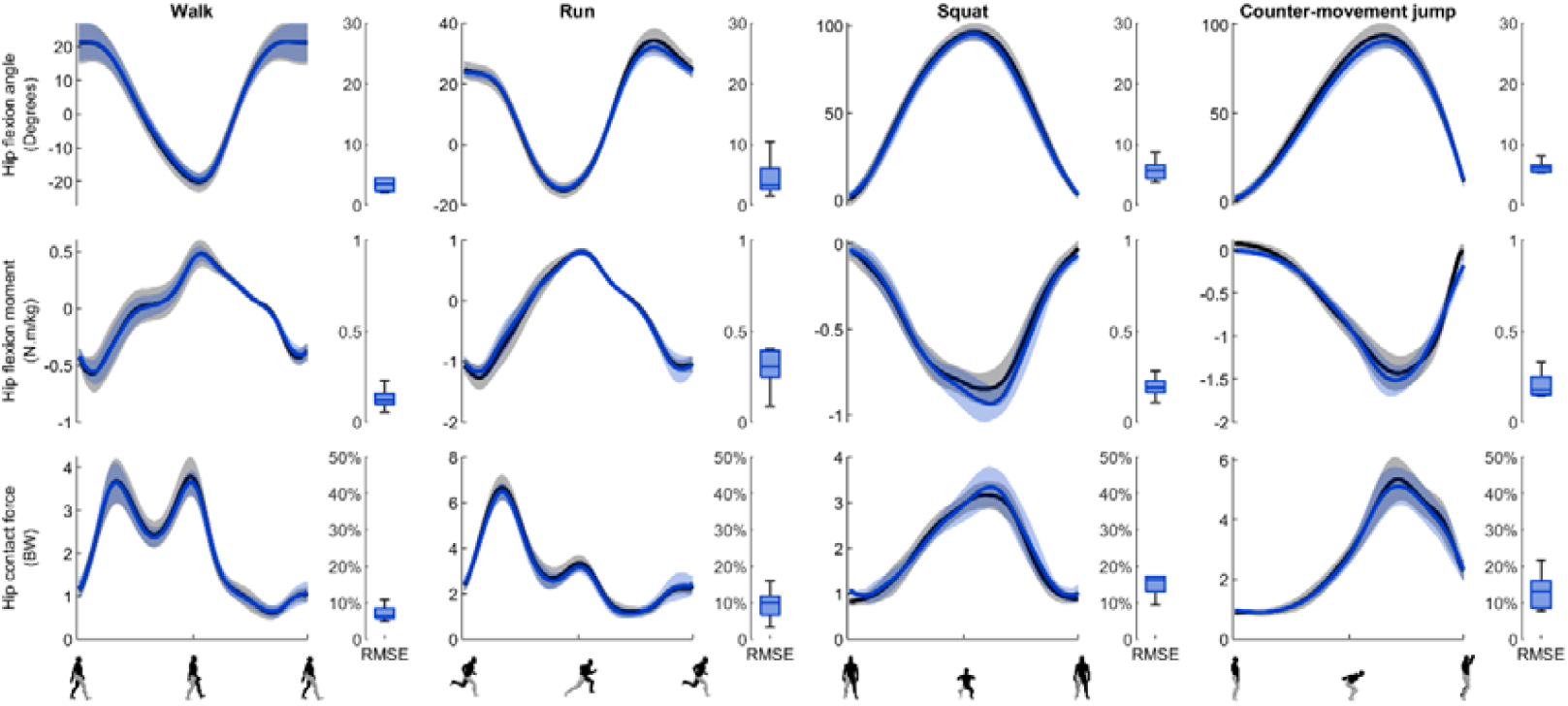
Neuromusculoskeletal model (NMS) estimations and physics-informed neural network (PINN) comparison. We compared our PINN predictions (blue) to a gold-standard electromyography-informed NMS model (black), using leave-one-subject-out cross-validation. The PINN well predicted sagittal plane joint angles (top-row), joint moments (middle row), and hip contact force magnitude (bottom row) across four distinct tasks. Mean (solid lines) and 95% confidence intervals (shaded region) are shown.

### Responsiveness to changes in peak hip contact force during walking

Beyond accuracy, the PINN must also be responsive to small changes in clinically important biomarkers to be a useful clinical tool. In the case of hip osteoarthritis, lower than normal magnitude and more concentrated regional hip articular loading have been identified as potential contributors to suboptimal cartilage mechanobiology, leading to degeneration^38,39^. Even small changes (e.g., +5%) in hip contact force magnitude and direction achieved via gait modification may alter resulting cartilage mechanobiology for therapeutic benefit (i.e., promote positive tissue adaptation) ^40^. As a measure of clinical utility, we assessed the responsiveness of the PINN to changes in peak hip contact force during walking (Fig 3). For each person, the mean stance phase peak hip contact force was identified and compiled according to gait condition (speed, incline, decline). Across all gait conditions, Bland-Altman analysis was used to compare 1^st^ peak hip contact force between NMS and PINN (Fig 3). The NMS showed an increase in the magnitude of the 1^st^ and 2^nd^ peak in hip contact force as walking speed increased. The PINN underpredicted the commensurate changes in the magnitude of the 1^st^ peak in hip contact force, with negligible bias -0.09 BW (LOA: -0.81 to 0.63 BW) and R^2^ of 0.76. For inclined walking conditions, NMS showed a decrease in the magnitude of the 1^st^ peak in hip contact force, while the declined walking conditions showed and increase in 1^st^ peak in hip contact force. The PINN had a matching response with negligible bias (inclined: -0.08 BW, LOA: -0.69 to 0.52; declined: -0.13 BW, LOA: -1.08 to 0.82). Overall, the PINN demonstrated the correct response to increases in the magnitude of the hip contact force, although the slope of the regression line was less than 1 indicating underprediction of high magnitude hip contact force and overprediction of low magnitude hip contact force.

**Fig. 3.**
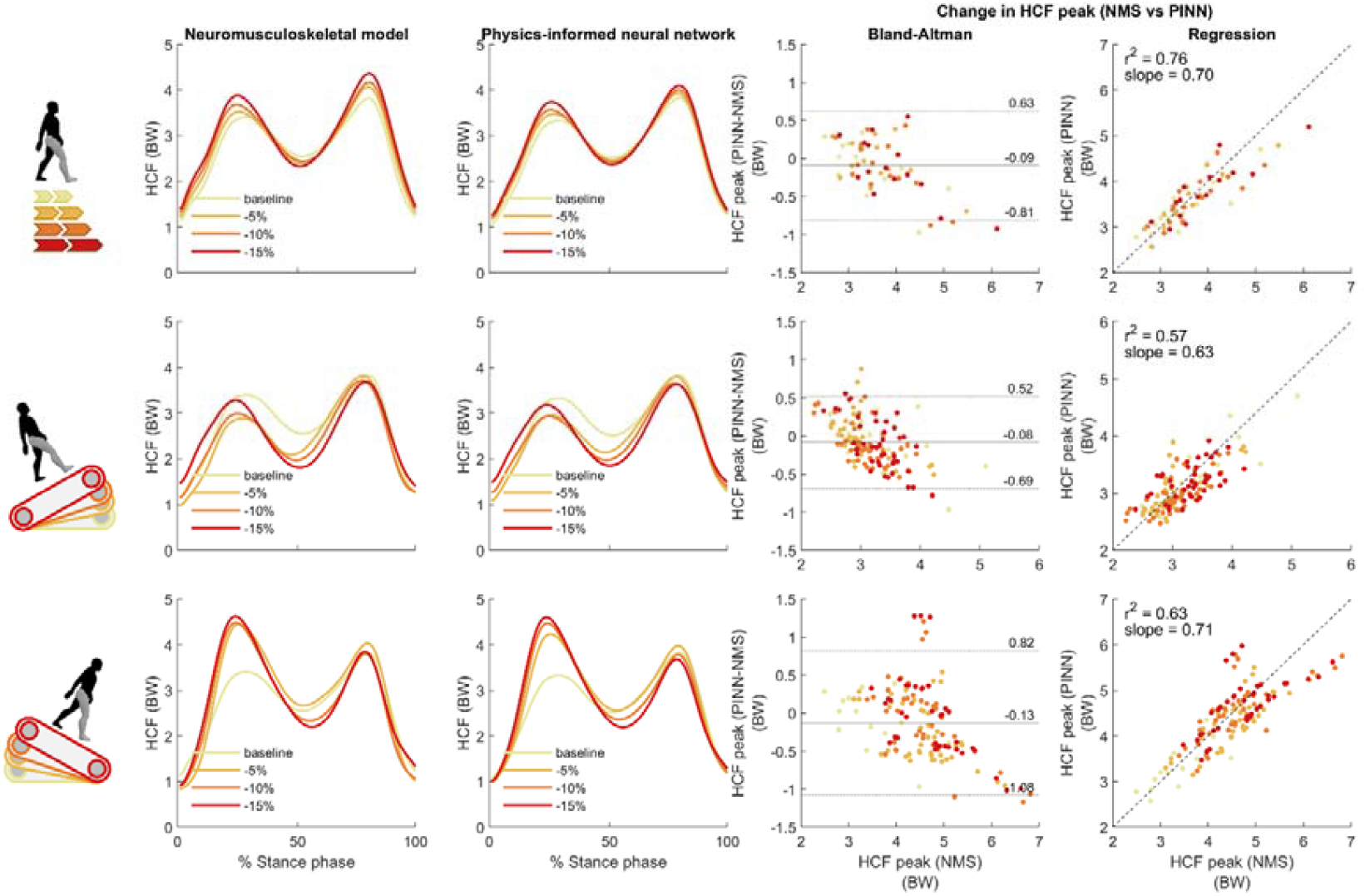
Comparison of NMS and PINN hip contact force predictions for different speeds and slopes. Mean NMS estimated (1^st^ column) and PINN predicted (2^nd^ column) hip contact forces are shown for increasing speeds (top row), incline gradient (middle row), and decline gradient (bottom row). Peak hip contact force was compared between NMS and PINN using Bland-Altman analysis (3^rd^ column) and linear regression (4^th^ column). Across all walking conditions, the PINN correctly predicted increases to 1^st^ peak hip contact force.

### Prediction of hip biomechanics during gait modification using a smart garment

Regarding internal biomechanical variables, NN have been suggested as a feasible alternative to laboratory-based data collection and modelling, though no study has demonstrated accurate real-time predictions using wearable technology. To demonstrate the feasibility of a wearable system for clinical, home, or field use, we used the PINN to estimate joint angles, moments, muscle forces, and hip contact forces from data acquired from a custom smart garment. The smart garment consisted of tight-fitting mid-thigh length shorts with embedded electronics (Supplementary Fig 1). The smart shorts contained three IMU (9-axis, 100 Hz), positioned over the sacrum and on the lateral side of each thigh. Four pairs of textile-electrodes were embedded over the gluteus maximus, gluteus medius, vastus lateralis, and biceps femoris. After onboard filtering, EMG linear envelopes from each muscle were down-sampled to match IMU sampling rate. Data from all sensors were synced, real-time streamed to a host computer, and used as input to the PINN. The smart shorts allowed prediction and visualisation of hip contact forces with a total delay between acquisition and prediction <40ms, enabling real-time biofeedback. We recorded data using smart shorts from a single participant (male, 25-30 years) who was asked to walk normally (preferred walking speed: 1.2 m/s), and with increased step width, increased step length, and anteriorly tilted pelvis. Data were recorded and analysed for 6 to 10 gait cycles per condition. For the increased step width condition (Fig 4, 1^st^ column), the PINN predicted lower peak hip adduction angle (1.3±0.6°), peak hip extension moment (0.06±0.04 Nm/kg), and 1^st^ peak hip contact force (0.32±0.11 BW) during the loading phase. For the increased step length condition (2^nd^ column), the PINN predicted greater peak hip flexion angle (4.1±0.8°) during toe-off and loading phase, greater peak hip extension moment (0.05±0.05 N.m/kg), and greater 1^st^ peak hip contact force (0.24±0.22 BW). When the participant was asked to anteriorly tilt their pelvis, the PINN predicted greater peak hip flexion angles with reduced hip extension during push-off (7.1±0.4°). The reduction in hip extension was accompanied by greatly reduced peak hip extension moment (0.22±0.03 N.m/kg) and 2^nd^ peak hip contact force (0.48±0.21 BW). When the hip contact force vector was projected onto the acetabulum, anteriorly tilting the pelvis was found to move the average position of loading backward thereby reducing loading to the anterior-superior portion of the acetabulum (Fig 4, right).

**Fig. 4.**
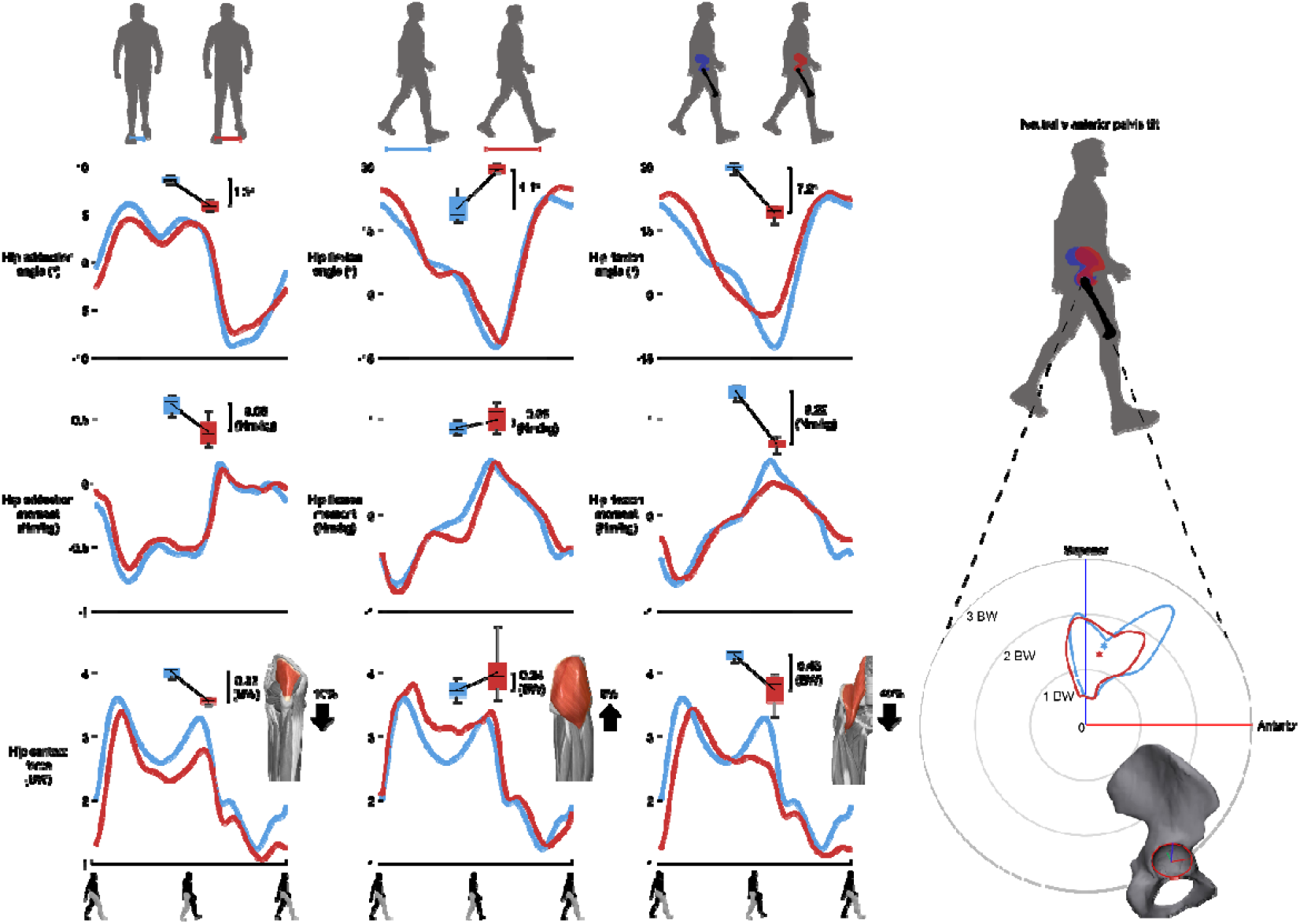
PINN predictions during gait modifications using a custom smart garment. Data recorded using a custom smart garment was used to predict hip angles (top row), hip moments (middle row), and hip contact force (bottom row). Predictions are shown for regular walking (blue) and compared to modified walking (red), for increased step width (1st column), increased step length (2nd column), and anteriorly tilted pelvis (3rd column). Walking conditions were analysed at regions of interest (shaded region) for which peak values are shown (box plots) with mean difference shown. The primary muscle responsible for differences in hip contact force (bottom row) is displayed, showing the mean change in muscle force from baseline condition. For the anteriorly tilted pelvis condition, the hip contact force vector was projected onto the acetabulum and is shown in acetabular coordinates (right). Shaded regions indicate discrete comparison, for which box plots show differences between conditions.

**Fig. 5.**
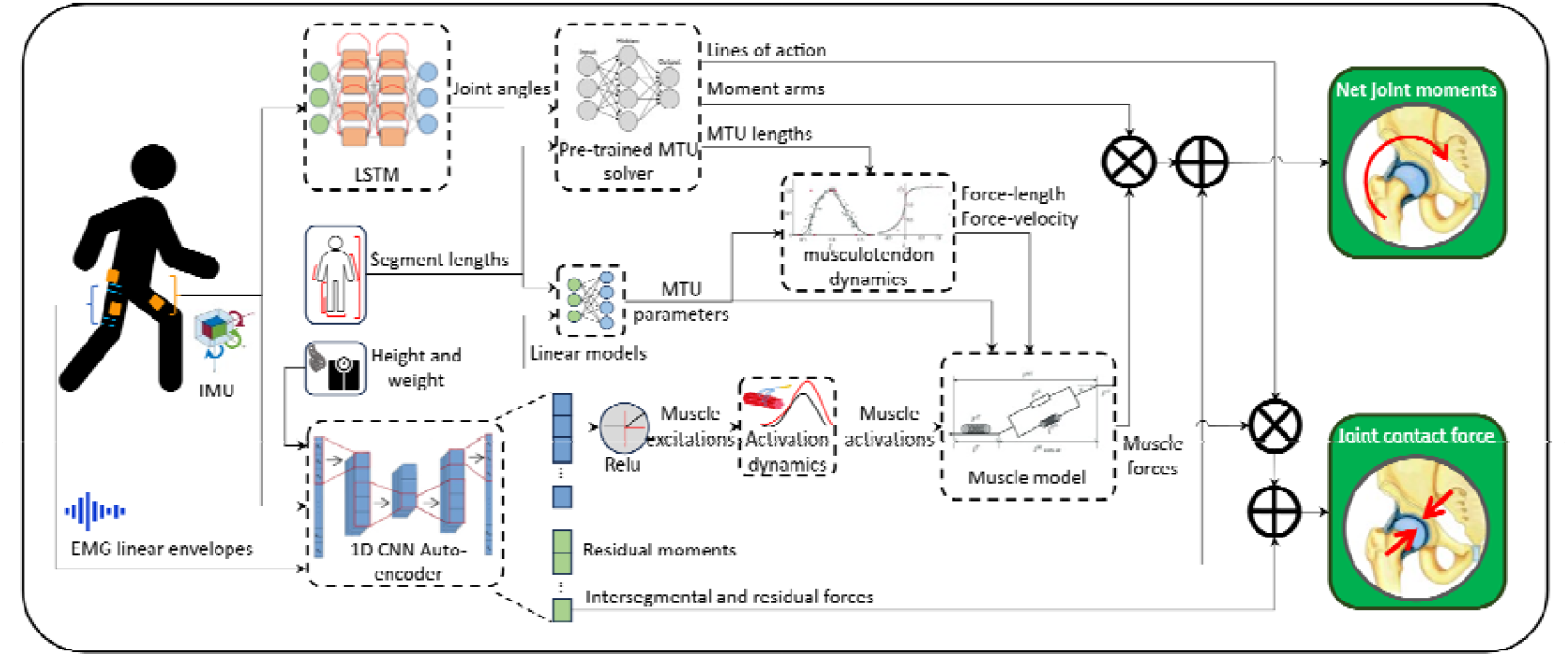
Overview of PINN architecture. Pre-processed data (rotated, normalised) from 3 IMU and 4 EMG are input to the PINN. The IMU linear accelerations and angular velocities are input to a LSTM model to predict joint angles. Predicted joint angles and estimated segment lengths for an individual are input to a pre-trained MTU kinematics solver that predicts MTU lengths, moment arms, and muscle lines of action. The MTU lengths are converted to normalised muscle fibre lengths using statistical estimates for MTU optimal fibre lengths and tendon slack lengths for each muscle. The normalised muscle fibre lengths are then used to compute force-length and force-velocity components for the muscle model. The EMG linear envelopes, IMU linear accelerations and participant height and weight are input to 1-dimensional CNN autoencoder to predict 25 muscle excitations, intersegmental forces, and residual forces and moments. The muscle excitations are passed through a rectified linear unit with a maximum value of 1, to bound excitations between 0 and 1. The bounded muscle excitations are then input to a critically damped filter and a non-linear scaler to produce muscle activations. Muscle activations, muscle force-length and force-velocity components, muscle pennation angles and estimated maximum isometric muscle forces are input to a Hill-type muscle model with a rigid tendon to compute muscle forces. Each muscle’s force is then multiplied by its instantaneous moment arm for hip flexion and adduction, and summed with residual moments to predict the net moment at the hip. Each muscle’s force is also multiplied by instantaneous line of action unit vectors and summed for each axis with intersegmental forces and residuals forces to predict hip contact force.

Muscle forces were computed within the PINN using a validated Hill-type muscle model. The changes to hip moments and hip contact force across walking conditions were explained by changes to muscle excitations and contraction dynamics that affect force output. For increased step-width, the PINN predicted reduced muscle excitation of the primary hip abductors (gluteus medius), resulting in lower gluteus medius force (∼10%) and lower peak hip adduction moments and hip contact force. For the increased step length position, increased hip extension moments and 1^st^ peak hip contact force were explained by increased excitation of the primary hip extensor (gluteus maximus) which produced ∼8% more force during early stance. Reduced iliopsoas force (∼40%) was the primary contributor to reduced hip flexion moments, reduced 2^nd^ peak hip contact force, and reduced anterior acetabular forces during walking with an anteriorly tilted pelvis. The changes to hip biomechanics elucidated by gait modification were consistent with underpinning neurophysiology and with findings from previous studies^40-44^, suggesting the PINN appropriately predicted muscle forces to account for changes in kinematics and kinetics during gait modification.

## Discussion

We developed a PINN to estimate clinically relevant biomechanics from wearable sensors and trained it to estimate mechanical biomarkers of interest during a gait retraining intervention for hip osteoarthritis. The PINN produced accurate predictions of hip angles, moments, muscle forces, and joint forces using only three IMU and four EMG, across multiple tasks, and in people not included in the training data. The PINN demonstrated responsiveness to changes in a clinical biomarker (hip contact force), a necessary characteristic for clinical utility. When paired with novel smart shorts, the PINN could predict hip biomechanics in real-time during a gait modification exercise and provide useful insights into the concomitant changes to hip contact force. The demonstrated smart garment system (Supplementary Fig 1) meets international demand for novel cost-effective health technologies^16,17^ to estimate and monitor biomarkers of tissue health in rehabilitation, disease management, sports performance, and injury prevention contexts^21^.

The capability of the developed PINN to estimate joint loads addresses a major scientific gap that has hindered the adoption of biomechanical interventions for musculoskeletal tissue disease^7^. For osteoarthritis, gait modification has shown some benefit to symptoms and mechanical biomarkers for the knee^45^ and hip^40^ but is not a recommended treatment due to a lack of high-quality evidence linking modification of cartilage loading with changes in symptoms and cartilage health^13^. The developed smart garment may enable verification of joint loading effects during gait retraining and provide direct biofeedback to users for step-by-step optimisation of biomechanical targets. The smart garment may also empower researchers with a tool for large-scale clinical trials of gait retraining in natural environments, a necessary step to assess treatment efficacy and update clinical practice guidelines^46^. As the PINN embeds known neural activation and muscle contractile behaviour that are common to the modelling of all muscles and joints^33^, it could be adopted for monitoring mechanobiology across life spans, populations, and conditions. For example, the PINN could improve rehabilitation outcomes of Achilles tendinopathy, for which the identification of optimal exercise type, intensity, and dose has long been hindered by the inability to measure Achilles forces/strains during therapy^47^. The use of the PINN and smart garment for management of musculoskeletal conditions is also supported by the potential cost-savings to health systems. Whereas typical motion capture systems for biomechanical analysis cost

>$US 1 million and require hours of analysis by expert staff, the total cost of parts for the smart garment prototype was <$US 1,000, and the system can be self-operated without technical expertise. Smart garments may also reduce the need for clinical visits and improve access for remote communities through technology-assisted telerehabilitation^48^.

Our results support the continued development of PINN and smart garments for monitoring of hip mechanics during gait modification, which has been identified as a potential disease modifying intervention^13,15,49^. On level ground, the PINN responded in a physiologically plausible way to gait modifications recorded from a smart garment (Fig 4), even though no data from gait modifications were used to train the model. Our results showed a correct response (e.g., increase or decrease) in peak hip contact forces during changes in speed and slope (Fig 3), though there was large variability in the magnitude of change. This variability may be explained by insufficient information in the input signals (i.e., little difference in thigh accelerations and angular velocities) and addressed *via* the addition of an IMU to the trunk to provide information of upper body dynamics, and to the feet to better detect changes in speed and slope. The required precision of monitoring tools such as smart shorts for osteoarthritis management is currently unknown. Small changes in joint loading (∼5%) may elicit meaningful responses in cartilage remodelling^9^ but clinically meaningful changes that connect joint load magnitude, direction, and cumulative load to optimal tissue-level mechanics (i.e., cartilage stresses and strains) have not yet been established^50^. By using the smart shorts to measure joint-level mechanics in natural environments, predictions may be further analysed using patient-specific tissue models (such as those derived from medical imaging)^51^ to model tissue mechanics and enable identification of optimal loading patterns. Future research may combine smart garment monitoring with gait modification interventions to establish clinically meaningful changes in hip mechanics, improve PINN training data size and diversity, inform sensor quantity and location, optimise garment design to meet consumer needs (e.g., cost, ease-of-use, comfort), and adapt the PINN for other musculoskeletal conditions (e.g., Achilles tendinopathy, osteoporosis).

Previous studies have demonstrated estimation of joint angles and moments from recordings of IMU data, though few have demonstrated real-time use. Our model improved upon previous work, estimating hip angles with high accuracy across multiple tasks (Fig 2). Whereas previous studies either allow natural variation in IMU placement^52^, or augment data to randomise IMU orientations^53^, we performed a static calibration that rotates all IMU accelerations and angular velocities to a common reference frame^18^. In the case of static calibration, the consistency of input data may improve learning of NN, for which gravitational accelerations align with a common axis, and gyroscopic rotations align with the anatomical axes of body segments. Hip moments and forces were estimated with higher percentage errors than joint angles which is likely due to the limited information on full-body dynamics provided by only three IMU. Whereas estimating joint angles can achieved by understanding the motion of articulating body segments, estimating joint moments and forces requires knowledge of applied forces on the body or accurate estimation of many muscle forces and moment arms. Nonetheless, the accuracy of the PINN to estimate hip moments was similar to previous work that used more input data (i.e., full-body motion)^54^, and with computationally expensive physics-based approaches that solve muscle forces and moments from full-body motion^27^.

A main barrier to large-scale adoption of NN methods in healthcare is a lack of explainability, and the “black-box” nature of model learning^55^. By integrating elements of physics-based NMS models within the NN, predictions of the developed PINN can be further investigated. For example, changes in joint moments or forces can be explained by changes in moment arms and muscle forces, which in turn can be explained by changes to neural activation or muscle contraction dynamics. Predicted muscle excitations of the PINN can be verified against recorded EMG, and muscle contractile dynamics can be investigated and compared to experimental data^56^ or physics-based simulations^57^. Although we integrated known neural and muscle contractile behaviour, multiple simplifications were made. The muscle model assumed a rigid tendon to simplify force calculations and negate the need to equilibrate tendon strain at the first timepoint of each input sample. Muscle-tendon kinematics relied on a surrogate of linearly scaled musculoskeletal models^58^, for which best practice was followed regarding optimisation of initial muscle-tendon parameters^59,60^. Nonetheless, the PINN inherits the same limitations (i.e., inability to personalise muscle paths, moment arms, and joint motion) of simplified NMS models that do not account for patient-specific musculoskeletal and joint anatomical geometry^61^. However, advancements in automated bone and muscle segmentation from medical imaging^62^, or statistical shape models that estimate personalised geometry from anatomical landmarks^63^, could be integrated into the PINN for improved patient-specific predictions.

Overall, our findings suggest the developed PINN can accurately estimate clinically relevant biomechanics from wearable technologies. We demonstrated this capability through a novel smart garment, which could be deployed in clinical, home, workplace, and sporting environments for the acquisition of biomarkers relevant to tissue health and adaptation. The presented PINN was used to estimate hip biomechanics but may be adapted and generalised for personalised management or rehabilitation of a broad range of musculoskeletal diseases and injuries.

## Methods

### Overview

The PINN was designed as a surrogate to a gold-standard NMS model, and to operate in real-time, using sparse IMU and EMG, enabling simple setup and use in ecologically valid environments. The PINN addressed the problems of moving NMS models outside the laboratory, such as the reliance on high-fidelity laboratory-based systems, which in many cases cannot be measured outside the laboratory (e.g., ground reaction forces), and the requirement of numerous hours of data cleaning, labelling, and simulation. To achieve robust and physiologically plausible predictions, the PINN incorporated key elements of NMS models, such as muscle contraction dynamics, and enforces mechanical consistency between outputs (i.e., muscle forces and joint moments). To ensure generalisability of the PINN, a gold-standard laboratory-based dataset consisting of diverse motions was collected synchronously to the IMU and then cleaned, labelled, and modelled using validated physics-based NMS modelling.

### Study design and participants

In this cross-sectional proof-of-concept study, seventeen participants (76.7±14.7 kg, 1.75±0.11 m, 28.0±2.8 years, 5 females) were recruited from a healthy population in Australia. Inclusion criteria were age (18-45 years); self-reported ability to walk, run, jump, and squat without pain or discomfort; and body mass index (BMI) <35 kg/m^2^. Participants were excluded if they had neuromuscular, neurological, or cardiovascular disease or injury that would affect their ability to perform testing; any spinal or back pain; and conditions that are contraindications to exercise or require medical clearance. This study was approved by the Institutional Human Research Ethics Committee (GU ref no: 2022/762) and participants gave their written informed consent prior to testing.

### Experimental setup

Three-dimensional marker trajectories (200 Hz) from 58 retroreflective markers were recorded using a 10-camera motion capture system (Vicon Nexus, Oxford Metrics, UK). Ground reaction forces (1000 Hz) were collected bi-laterally using an instrumented treadmill (AMTI, USA) and ground-embedded force plates (AMTI, USA). Surface EMG (Cometa, Italy) were recorded at 2000 Hz from 14 lower limb muscles from the dominant side: gluteus maximus, gluteus medius, tensor fascia late, rectus femoris, sartorius, gracilis, vastus lateralis, vastus medialis, biceps femoris, semitendinosus, gastrocnemius lateralis, gastrocnemius medialis, soleus, and tibialis anterior following a standardised protocol^64^. Linear accelerations, angular velocities, and magnetic field vectors (100 Hz) were recorded using IMU (XSENS, Netherlands) placed on the pelvis and left and right thighs.

### Procedure

The EMG were recorded during maximal voluntary isometric contractions (MVIC) for later normalisation in NMS modelling. Each participant performed maximal effort hip flexion, hip extension, hip adduction, hip abduction, knee flexion, knee extension, ankle plantarflexion, ankle dorsiflexion, and combined hip flexion, hip external rotation, and knee flexion (sartorius contraction). Marker trajectories and IMU were recorded during a static trial (anatomical position) for model scaling and IMU calibration.

Participants performed treadmill walking, treadmill running, overground walking, squatting, countermovement jumping, forward jumping, side-step cutting, and a sit-to-stand task. Participants were first asked to walk on a treadmill at 0.9 m/s, and the speed was increased by 0.05 m/s every 30 s until the participant stated it matched what they believed to be their typical walking pace (baseline). After a further 2-minute familiarisation, biomechanical data were recorded for 30 s. The treadmill speed was then increased by 5, 10, and 15% of baseline and 30 s of data were recorded at each of these increments. Each participant then ran at twice the speed of their baseline walking speed and +5%, +10%, and 15% of baseline walking and running speeds. The participants were then given 5 minutes of rest after which they performed walking trials again at baseline, +5%, and +10% speeds for both incline and decline (5%, 10%, and 15% grade) with 5 minutes of rest between each grade.

The participants then performed overground walking (self-selected pace), followed by body-weight squats, split-squats with both left and right legs forward, countermovement jumps, forward jumps, sit-to-stand, and running with a side-step cut. Eight repetitions were completed per task. The posture (e.g., foot position, squat depth), intensity (e.g., jump effort), and movement duration were not controlled to ensure the dataset included natural variation.

### Musculoskeletal modelling

Raw C3D files from Vicon Nexus v2.9.1 (Oxford Metrics, UK) were processed using the MOtoNMS toolbox^65^ in MATLAB (MathWorks, USA). Marker trajectories and ground reaction forces were low-pass filtered (6 Hz) using a 2^nd^ order, dual-pass, zero-lag Butterworth filter. To calculate muscle excitations, EMG were band-pass filtered (30–300 Hz), full-wave rectified, and low-pass filtered (6 Hz) with a 2^nd^ order, dual-pass, zero-lag Butterworth filter, then amplitude normalised to the maximum filtered value from all MVIC and dynamic trials.

A generic full-body OpenSim model^66^ was scaled using anatomical landmarks from each participant. The hip joint centres, with respect to pelvis markers, were estimated from each participant’s leg length, measured from markers placed on the anterior superior iliac spine, medial femoral condyle, and medial malleoli^67^. The knee joint centre was defined as the midpoint between markers on the femoral condyles, the ankle joint centre was defined as the midpoint of the medial and lateral malleoli plus an inferior offset of 2.7% of shank length^68^. After scaling, tendon slack and optimal fibre lengths for each muscle-tendon unit were optimised to ensure physiological operating ranges^59^, and the maximal isometric force of each muscle was estimated from the participant’s height and weight^60^. OpenSim^32^ inverse kinematics, inverse dynamics, and muscle analysis tools were used to calculate joint angles, joint moments, and muscle-tendon kinematics, respectively, and the process was streamlined using the Batch-OpenSim-Processing-Toolbox (BOPS) ^69^. For each subject, the calibrated EMG-informed neuromusculoskeletal modelling (CEINMS) toolbox^33^ was used to first optimise neuromusculoskeletal muscle-tendon parameters and then determine muscle-tendon unit dynamics. During optimisation, maximal isometric strength, optimal muscle fibre length, tendon slack length, and excitation to neural activation filter coefficients and non-linear shape factor were modified to reduce the difference between muscle-driven and experimental moments using a hybrid EMG-driven and synergy-based calibration^70^. For each trial, the calibrated NMS model was executed in EMG-assisted mode, which allows small adjustments to muscle excitations, and synthesis of non-measured excitations to match experimentally measured moments:

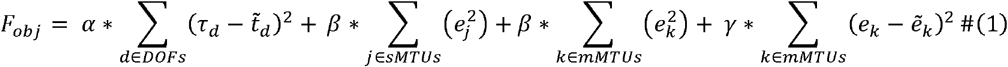

where, *τ*_*d*_, is the moment at the *d*^th^ joint estimated by CEINMS, 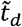, is the experimental moment at the *d*^th^ joint determined through inverse dynamics analysis, *e*_*j*_, is the estimated excitation for the *j*^th^ non-measured MTU in the model, *e*_*k*_, is the estimated excitation for the *k*^*th*^ measured measured MTU in the model, and 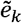 is the experimental excitation for the *k*^th^ MTU in the model. Finally, the muscle driven solution was input to OpenSim joint reaction analysis to estimate three-dimensional hip contact force. Muscle excitations from all 14 recorded EMG were used for both calibration and execution in CEINMS. An electro-mechanical delay of 38ms was used in CEINMS which is within reported guidelines^71^ and coincides with the delay induced by online filters used for the PINN and smart garment^72^. Motion trials used for CEINMS calibration were not then used for model execution.

### Neural network data pre-processing

During quiet upright stance, the on-board Kalman filter for the IMU (XSENS, Netherlands) was used to calculate the global heading using the IMU placed on the pelvis. For the present study, the local z-axis of the pelvis IMU was positioned facing forward, thus the heading was calculated according to:

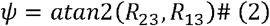

where, *Ψ*, is the yaw angle denoting the forward-facing direction,*atan*2, is the two-argument arctangent function, (*R*_23_,*R*_13_, are the rotation matrix components that represent the IMU frame z-axis rotation in the global x-y plane. The anatomical axes (x-axis = anterior-posterior, z-axis = superior-inferior, y-axis = medial-lateral) were then approximated according to:

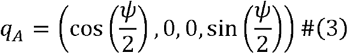

where,*q* _*A*_, is the quaternion representing the anatomical frame. For each IMU (pelvis, right thigh, and left thigh) raw accelerometer, gyroscope, and magnetometer data from dynamic trials were rotated to the anatomical frame according to:

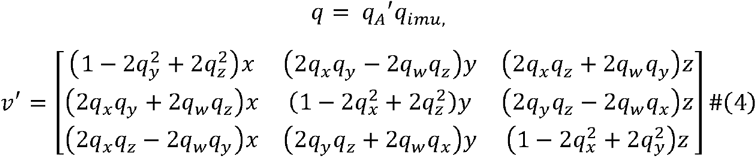

where, *v*^′^, is rotated signal vector and, *q*, is the offset between offset between the static orientation of an IMU (*q*_*imu*_) and the anatomical frame.

The EMG linear envelopes of four hip-spanning muscles (gluteus maximus, gluteus medius, rectus femoris, and biceps femoris) were selected for input to the neural network. The EMG were filtered in the same manner as for NMS, though only a single pass was performed to enable future real-time use. The resulting linear envelopes were amplitude normalised to the maximum value recorded during walking at a self-selected pace, which can be easily performed in practical applications.

The IMU and EMG data were concatenated, and a sliding window method was used to generate 300 ms samples (30 frames) for NN training. For each person, static input data were also generated, and included each participant’s height and mass, body segment scaling factors (size of pelvis, thigh and tibia relative to unscaled OpenSim model).

### Neural network architecture

The neural network consisted of three sub-networks (Fig 2): a kinematics estimator, MTU kinematics estimator, and an autoencoder model that generates both muscle excitations and intersegmental force estimations (NMS solver). The kinematics estimator used pelvis and thigh IMU data to estimate hip flexion, hip adduction, hip rotation, and knee flexion angles. The MTU kinematics estimator was a previously trained and validated feed-forward NN that estimates muscle-tendon lengths, moment arms, and lines of action of all hip-spanning muscles from the predicted joint angles and anatomical factors^58^. The NMS solver used the four EMG linear envelopes, and IMU linear acceleration and angular velocities, to generate muscle excitations for 25 hip spanning muscles that were bounded between 0 and 1, in addition to 3-axis intersegmental forces in the pelvic coordinate system. The NMS models were calibrated to each individual, whereas the PINN had to learn a generalisable solution to estimate muscle excitations. Consequently, the NMS solver predicted residual hip forces and moments to account for discrepancies between the PINN’s muscle driven solution and the ground-truth moments and forces from the NMS model.

The PINN-generated muscle excitations were used to compute muscle forces at each timestep according to:

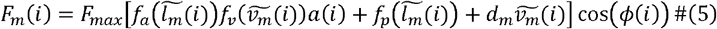

Where, *i*, is a discrete time point, *F*_*m*_ (*i*), is the muscle force, *F*_*max*_, is the maximum isometric muscle force, *f*_*a*_, is the active force-length component, 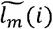, is the normalised muscle fibre l e n g th, *f*_*v*_, is the force-velocity component, 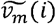, is the normalised muscle fibre velocity, *a*(*i*) is the muscle activation, *f*_*v*_, is the passive force length component, 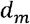, is the muscle-damping coefficient (set to 0.1), *ϕ* (*i*) is the muscle pennation angle. A rigid tendon model was used to enable rapid calculation of muscle fibre lengths:

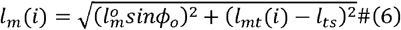

where, *l*_*m*_ (*i*), is the normalised muscle fibre length, 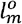, is the optimal muscle fibre length, *l*_*mt*_, is the muscle-tendon length, and *l*_*ts*_, is the tendon slack length. Muscle activations were calculated from muscle excitations by modelling the muscle twitch response:

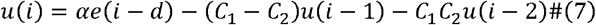

Where, *u*(*i*), is the neural activation, *e*(*i*), is the muscle excitation, *α*, is the muscle gain, and *C*_1_ and *C*_2_, are recursive coefficients. The neural activation is the converted to muscle activation using the non-linear relationship:

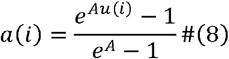

where, *a*(*i*), is the muscle activation, and *A*, is the on-linear shape factor constrained between -3 and 0.

To enable ease of use of the PINN, each participant’s initial muscle-tendon unit properties were calculated from their mass, height, and lower-limb anatomical scale factors using multivariate linear models. The optimal muscle fibre and tendon slack lengths were estimated with error (r^2^>0.997, RMSE: 0.18±0.15mm and r^2^>0.983, RMSE: 0.23±0.16mm) compared to OpenSim models that were optimised to preserve physiological operating ranges^59^. Maximum isometric muscle forces were estimated with negligible error (r^2^>0.999, RMSE: 3.25±2.22N) compared to OpenSim models with forces adjusted to account for height and mass^60^. The linear models for all MTU parameters were vectorised and integrated as a single matrix operation within the PINN. Equation coefficients and fitting errors are detailed in Supplementary Table 1.

Joint moments were calculated within the PINN as the sum of predicted muscle forces multiplied by their respective moment arms at each timestep and the additional predicted residual moments. Three-dimensional hip contact force was calculated by multiplying each muscle’s scalar force with its corresponding line-of-action unit vector, summing the force along each axis, and adding the predicted intersegmental forces and residual forces. The kinematics estimator was a multi-layer long short-term memory (LSTM) network, and the NMS solver was a 1-dimensional convolutional NN autoencoder. Whereas the kinematics and kinetics estimators were trained to predict outputs from OpenSim (angles, moments, intersegmental forces), the excitation generator was not provided any training labels. As the muscle excitations from CEINMS are specific to a calibrated model (i.e., adjusted MTU parameters), which is infeasible in practical settings, the activation generator was instead trained to produce muscle activations that minimise the difference in resulting muscle forces and hip contact forces.

The NN were developed, trained, and evaluated in TensorFlow (v2.16). The NN were trained using a combination of data-driven loss functions (i.e., predicted vs ground-truth) and additional penalties that encourage mechanical consistency between muscle forces, joint moments, and joint forces, and enforce known neural and contractile dynamics. The standard data-driven regression loss function was modified to reduce differences in the 1^st^ (angles, moments, and forces) and 2^nd^ (angles only) derivatives of predicted and ground truth data, resulting in physiological (e.g., smooth) outputs:

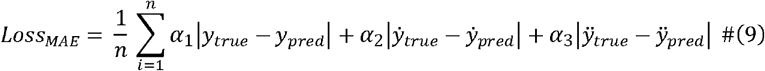

where, *y*_*true*_ are training labels from OpenSim and CEINMS, *y*_*pred*_ are the predicted joint angles, joint moments, intersegmental forces, muscle forces, and hip contact forces, where (.) and (^..^) indicate first and second time derivative, respectively, *M*_*pred*_ are the predicted joint moments from the kinetics estimator, *mF*_*pred*_ are the are the muscle forces calculated from the generate activations, *m*_*r*_ are the muscle moment arms, *n* is the number of outputs, *m* is the number of muscles, and *α*_1_, *α*_2_, and *α*_3_, are pre-calculated coefficients that provide equal weightings to the total loss (i.e., adjusting for larger values of derivatives). The NN model hyper-parameters (i.e., number of layers, number of LSTM cells, regularisation coefficients, learning rate, batch-size) were selected using Bayesian and HyperBand optimisation with a train-test split of 3:1 (Supplementary Table 2).

To encourage physiologically plausible muscle excitations, additional penalties were added to the NMS solver that generates muscle excitations:

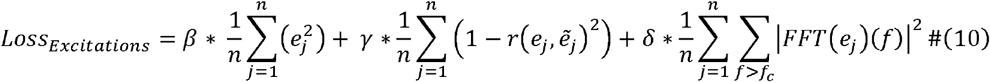

where, *β*, is the same as for NMS models, and penalises total muscle excitations, *r*, is the correlation between predicted excitations, *e*, and measured EMG linear envelopes, 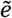, and *γ*, is weighting applied to poor correlation between *e* and 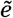. The excitation penalty of the NMS model (Eq. 1) was modified to account for the different EMG normalisation method for PINN and thus penalises poor correlation instead of difference. Finally, a Fourier loss was applied to all predicted excitations to penalise high frequency non-physiological excitations. The fast-Fourier transform, *FFT*, for each excitation is computed and the power spectral density for all frequencies, *f*, above a cut-off frequency, *f*_*c*_, are summed. *δ*, is the weighting applied to the Fourier loss. The cut-off frequency was set to 6Hz. Vanishing gradients that could lead to erroneous flat excitations during training, were prevented *via* a soft-constraint that penalised excitations beyond acceptable ranges (0 to 1) and with zero variance, before being clipped between 0 and 1 when input to the muscle model.

Whilst optimal fibre lengths, tendon slack lengths, and maximum isometric muscle forces were estimated from anthropometrics, small adjustments (±15%) were allowed during training to account for common alterations that may occur during EMG-informed calibration. Changes to MTU kinematics were constrained by applying a penalty to normalised fibre lengths outside physiological ranges:

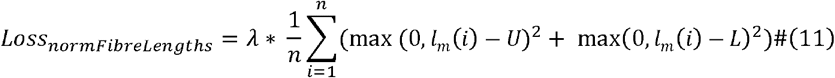

Where, *l*_*m*_ (*i*), is the normalised muscle fibre length, *U*, is the upper limit for fibre length (set to 1.4) and *L*, is the lower limit for fibre length (set to 0.6), and, is weighting applied to the loss. The PINN included residual moments and forces to account for errors in muscle force prediction, and effects of small differences in moment arms and lines of action that occur due to errors in joint angle estimation. To prevent the PINN from over-using residual force and moments and consequently producing poor muscle force predictions, an additional penalty was applied to penalise residuals:

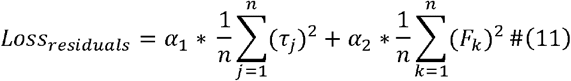

where, *τ*, are residual moments for the *j*^*th*^ hip degrees of freedom, *F*, are residual forces acting on each axis, *n*, and *α*_1_, *α*_2_ are penalty weightings.

### Neural network training

The PINN was trained using NMS data in 3 steps. First, the weights of the NMS solver were kept constant, and the joint angle estimator was pre-trained to avoid non-physiological MTU kinematics that prevent the excitation generator converging. Second, the angle estimator weights were frozen, and the NMS solver was trained, resulting in predictions of muscle forces, net joint moments, intersegmental forces, and hip contact force. Finally, the angle estimator weights were unfrozen, and the PINN was further trained to allow feedback from muscle force estimation to joint angle estimation (i.e., hip contact force and net moments are influenced by MTU kinematics that are a function of joint angles).

After training, the PINN can be fine-tuned for an individual without the need for labelled NMS data. Individual-specific calibration is achieved by first freezing the weights of the angle estimator and NMS solver of a trained model. Weightings for MTU parameters (optimal fibre length, tendon slack length, and maximum isometric force) and muscle activation parameters (shape factor and recursive coefficients) are unlocked and allowed to adjust within predefined ranges (Supplementary Table 3). The trained PINN can then be used without reference data, for which the loss function minimises the residual moments and forces by improving the muscle-driven solution.

### Neural network validation

The predicted angles, moments, muscle forces, and hip contact forces were compared using leave-one-subject-out cross-validation. We compared the coefficient of determination, root-mean-square-error (RMSE), and normalised RMSE (nRMSE). For directional values (angles, moments, 3-axis forces) nRMSE was normalised to range. For scalar variables (muscle forces and hip contact force magnitude) nRMSE was normalised to peak values. To as assess responsiveness of the PINN to detect changes in hip contact force elucidate by changes in gait, we compared the peak predicted and ground-truth hip contact force for all walking trials (baseline, fast, inclined, declined) using Bland-Altman and linear regression.

### Smart garment proof of concept

Custom smart shorts (Australian Centre for Precision Health and Technology) were used to record IMU and EMG to test the feasibility of using the PINN within a novel gait retraining technology for management of hip osteoarthritis. The smart shorts consisted of a network of custom-designed electronics, including miniaturised IMU sensors and EMG amplifiers^73^. Three IMU sensors were embedded in the shorts (sacrum, right thigh, and left thigh). Washable conductive textile dry electrodes (Electroskin GECKO, Nanoleq, Switzerland) were embedded to acquire EMG from four muscles of the right leg (gluteus maximus, gluteus medius, vastus lateralis, and biceps femoris) ^74^. Two bipolar electrodes were embedded per muscle, with a single ground electrode positioned at the left anterior superior iliac spine. Sensors were enclosed in custom-designed 3D printed housings made from polylactic acid (PLA) (Bambu Lab, China) and were easily removable from their respective 3D printed docks, which were affixed to the garment using an iron-on process. The docks were wired to a central HUB (sacrum), which powered the network of sensors, provided data communication *via* I2C protocol, and wirelessly streamed data to a remote receiver. The smart shorts IMU sensors outputted raw accelerations, angular velocities, and magnetic field vectors in addition to orientations computed via a Kalman filter (100 Hz). The smart shorts raw EMG (1000 Hz) were filtered (see neural networking pre-processing) to provide EMG linear envelopes (100 Hz).

A single participant donned the smart shorts and was asked to walk at a self-selected pace on a treadmill. During treadmill walking, the participant was also asked to walk with 3 separate modifications to their gait (longer steps, wider steps, and with an anteriorly tilted pelvis). The collected smart shorts data was used to predict joint angles, moments, muscle forces, and hip contact forces using the trained PINN. The predicted joint angles, moments, muscle forces, and hip contact force from each gait modification were compared to baseline.

## Data Availability

Data produced in the present study are available upon reasonable request to the authors

## Data Availability

Data produced in the present study are available upon reasonable request to the authors

## Acknowledgements

This study was supported by PhD Scholarship from Griffith University (B.C.); National Health and Medical Research Council (NHMRC) Investigator Grant Emerging Leadership Level 1 2017012 (L.D.); Discovery Early Career Research Award (DECRA) DE220101249 (D.S.); and Bionics Queensland Major Prize (B.C.)

