## Supplementary Fig 1 for "Real-time hip biomechanics from smart garments via a physics-informed neural network"

| **Supplementary Table 2.** Model architecture and hyperparameter tuning. | | | |
| --- | --- | --- | --- |
| **Hyperparameter** | **Minimum/Maximum or types** | **Increment/factor** | **Tuned** |
| *kinematics estimator (LSTM)* | | | |
| Number of layers | 1/3 | 1 | 2 |
| LSTM cells per layer | 32 | 512 | 128,64 |
| L2 regularisation | 0, 1e-5/0.1 | x10 | 0.01 |
| Temporal dropout fraction | 0/0.5 | 0.05 | 0.05 |
| Normalisation | None, Batch, Layer | -- | None |
| *NMS solver (1D-CNN autoencoder)* | | | |
| Number of encoder/decoder layers | 2/6 | 1 | 4 |
| Number of filters (1st layer)^†^ | 32/512 | x2 | 64 |
| Filter size | 3/9 | 2 | 3 |
| Temporal dropout fraction | 0/0.5 | 0.05 | 0.05 |
| Encoder normalisation | None, Batch, Layer | -- | Layer |
| Latent size | 4/32 | x2 | 8 |
| Increment refers to the step size between each tested hyperparameter value between minimum and maximum. When ‘x’ is present, the hyperparameter is increased successively by the provided factor from minimum to maximum (e.g., 0.001, 0.01, 0.1). Temporal dropout is a modified version of a dropout layer that drops entire feature maps (i.e., all timepoints) as opposed to a random fraction of the full tensor. ^†^The number of filters is halved in each successive encoder layer. The number of filters in the decoder layers are opposite to encoder (e.g., encoder: 128,64,32; decoder: 32,64,128). | | | |

| **Supplementary Table 3.** Initial values and ranges of muscle-tendon unit parameters for unsupervised subject-specific calibration. | | |
| --- | --- | --- |
| **Muscle-tendon parameter** | **Initial value** | **Range** |
| Maximum isometric strength | 1 | [0.75, 1.25] |
| Optimal muscle fibre length | 1 | [0.85, 1.15] |
| Tendon slack length | 1 | [0.85, 1.15] |
| Recursive coefficient (C1) | 0.495 | [0, 1] |
| Recursive coefficient (C2) | 0.505 | [0, 1] |
| Non-linear activation shape factor | -0.001 | [-3,-0.001] |
| Activation scale factor | 1 | [0.75, 1.25] |
| The initial values for muscle contractile properties (maximum isometric strength, optimal muscle fibre length, and tendon slack length) for each muscle were estimated from the participants height, mass, and body segment dimensions. Thus, the initial value and ranges for contractile properties are ratios relative to subject-specific estimated values. Recursive coefficients C1 and C2 were initialised with different values to avoid identical gradients during backpropagation. | | |

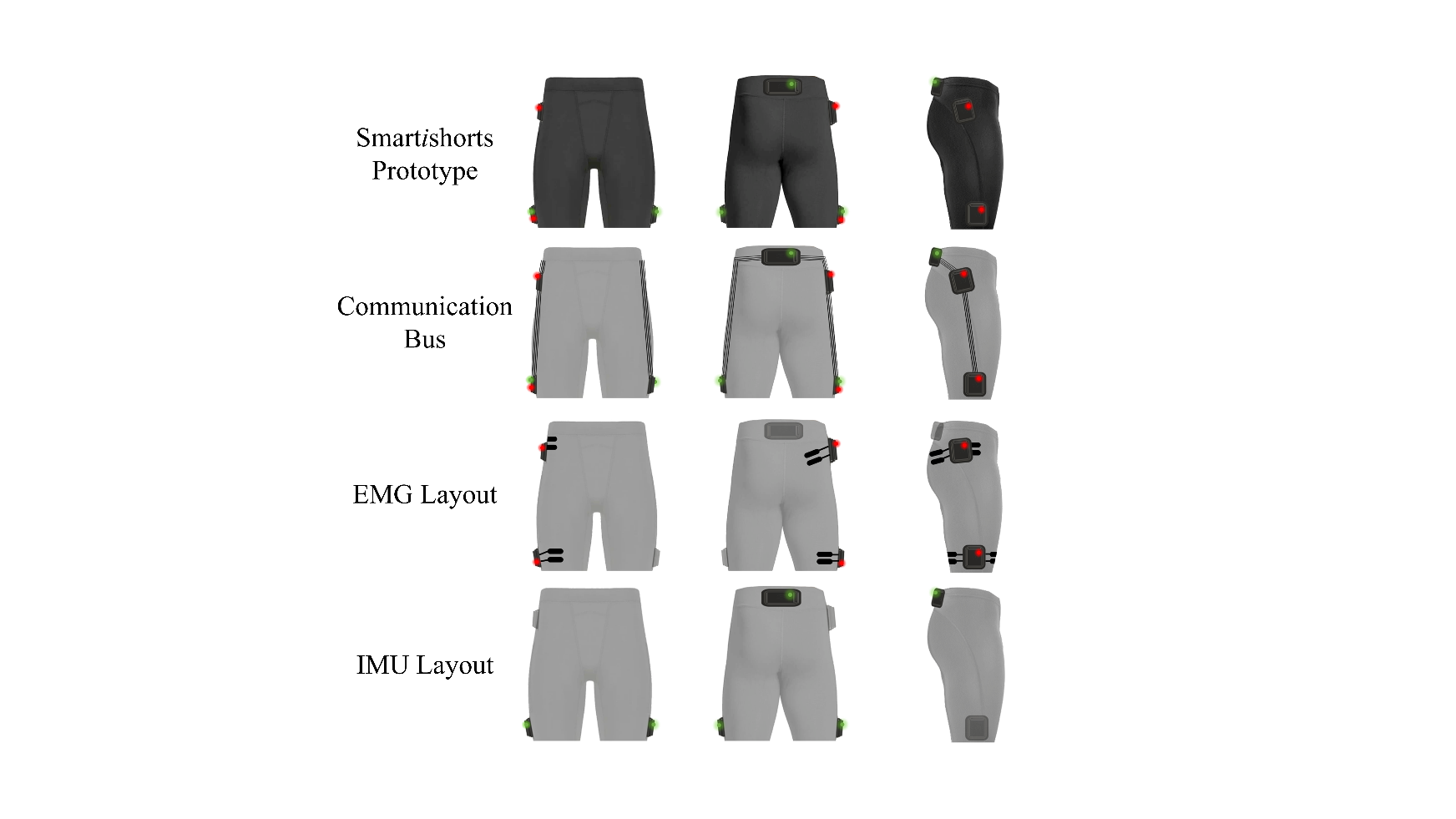

**Supplementary Fig 1 | Smart shorts prototype garment.** The Smart shorts prototype garment consisted of a network of custom-designed electronics, including miniaturised inertial measurement units (IMU) sensors and electromyography (EMG) amplifiers. Three IMU sensors were embedded in the shorts (4^th^ row: sacrum, right thigh, and left thigh). Washable conductive textile dry electrodes (Electroskin GECKO, Nanoleq, Switzerland) were embedded to acquire EMG from four muscles of the right leg (3^rd^ row: gluteus maximus, gluteus medius, vastus lateralis, and biceps femoris). Two bipolar electrodes were embedded per muscle (interelectrode distance = 20mm), with a single ground electrode positioned at the pelvis. Sensors were enclosed in custom-designed 3D printed housings made from polylactic acid (PLA) (Bambu Lab, China) and were easily removable from their respective 3D printed docks, which were affixed to the garment using a custom iron-on process. The docks were wired to a central HUB (sacrum), which powered the network of sensors, provided data communication via I2C protocol, and wirelessly streamed data to a remote receiver. The smart shorts IMU outputted raw accelerations, angular velocities, and magnetic field vectors in addition to orientations computed via a Kalman filter (100 Hz). The smart shorts raw EMG (1000 Hz) were filtered (see neural networking pre-processing) to provide EMG linear envelopes (100 Hz).
